# Women’s autonomy, neonatal, infant and under-five mortality in the Upper East Region of Ghana

**DOI:** 10.1101/2023.12.14.23299959

**Authors:** Fabian Sebastian Achana, Augustine Tanle, David Teye Doku

## Abstract

**Background:** Eight years to the set deadline for the 2030 SDGs, child mortality remains a major health challenge in Sub-Saharan Africa. Child survival is greatly influenced by household circumstances and mother’s healthcare choices. Notwithstanding tremendous investment in women empowerment in Ghana, there is limited empirical evidence on whether women’s autonomy translates into better child mortality outcomes.

**Objective:** To examine the association between women’s autonomy and neonatal, infant and under-five mortality in the Upper East Region of Ghana.

**Methods:** Data were obtained from a randomized cluster household survey among 15-49 years old women in seven districts in the Upper East Region. Data analysis was restricted to 3,243 women who reported ever having given birth. Based on Principal Component Analysis (PCA), we constructed an autonomy index categorized into least, moderate, and high autonomy based on responses to six questions regarding household decision-making. Bivariate and multivariate logistic regressions were used to assess the association of women’s autonomy status and mortality outcomes.

**Results:** Attaining secondary education or higher was significantly associated with infant mortality (adjusted odds ratio (aOR)= 0.39, CI= 0.16, 0.94) and under-five mortality (aOR= 0.39, CI= 0.18-0.87). Also, maternal age was significantly associated with neonatal, infant, and under-five mortality, while living in rural setting was significantly associated with lower risk of neonatal (aOR= 0.38, CI=0.19-0.75) and under-five (aOR= 0.63, CI= 0.48-0.83) mortality. However, we found that compared to women with least autonomy, infants of those with moderate autonomy (aOR= 1.76, CI 1.07-2.89) and high autonomy (aOR= 1.75; CI= 1.04- 2.93) were significantly more likely to die.

**Conclusion:** In this study setting, women’s autonomy was not predictive of child mortality. Interventions that aim to improve child mortality should pay attention to community and family level factors that promote increase utilization of essential early childhood interventions.

## Introduction

Globally, child mortality improved substantially over the years. It is estimated that 3.7% of children died before age five in 2021 compared to 8.3% in 2010 [1,2]. This notwithstanding, child mortality remains a major health challenge especially in Sub-Saharan Africa where 1 out of every 13 children die before age five compared to 1 in 189 in the developed nations. Sub-Saharan African (SSA) countries face a disproportionate burden of child mortality with all six countries presenting with the highest under-five mortality of approximately 100 deaths per 1,000 live births [3–5] . Within the continent, infant mortality ranges from 64 deaths per 1,000 live births in West Africa to 24 deaths per 1,000 live births in North Africa [6]. In Ghana, the latest available national data shows that neonatal mortality declined from 28 deaths per 1000 live births in 2015 to 17 deaths per 1000 live births in 2022. Similarly, under-five mortality declined from 60 deaths per 1000 live births in 2015 to 40 deaths per 1000 live births and infant mortality stood at 28 deaths per 1000 live births. Mortality during the first month (neonatal mortality) accounts for 61% of infant deaths and 43% of under-5 deaths [7].

However, there is wide disparities in the regional burden of child mortality, with the lowest rates of 47 deaths per 1,000 live births occurring in the Greater Accra region and the highest, “111 deaths per 1,000 live births” in the Northern region of Ghana. The Upper East region where this study was conducted has the fourth highest Under-5 mortality of 72 deaths per 1000 live births” following the Upper West regions’ “92 deaths per 1,000 live births”, and Ashanti regions’80 deaths per 1,000 live births [8]. Identifying the causes of deaths and promoting good child health in these regions with high mortality is key to achieving substantial overall reductions in child mortality in Ghana.

The healthcare system in the Upper East region has been described as fragile with weak referral care services and low availability of skilled staff, logistics, and medicines [9]. In addition, the region represents a typical patriarchal society, characterized by male dominance, restrictive female cultural norms and practices, early marriage and childbearing among women [10, 11]. Thus, gender inequalities are highly prevalent and skewed disproportionately against women [8].

Several biological, health system and demographic, socioeconomic and cultural factors have been implicated in the cause of child mortality in Ghana [8,12–14]. Women empowerment and autonomy has been recognized as an important predictor of maternal and child healthcare utilization, child survival and wellbeing [13, 15–21]. Over the years, substantial efforts have been made in Ghana to empower women and improve maternal and child health [8, 22]. These include provisions in the 1992 Constitution of Ghana especially Article 17 (1) and (2) which guarantees gender equity, nondiscrimination, and social justice [23]. Further to these constitutional provisions, Ghana is a signatory to several international treaties on women and children’s rights and committing to attaining international goals such as the sustainable development goals (SDGs). Other interventions include the implementation of the Health Sector Medium-Term Development Plan 2010-2013, the Under-five Child Health Policy (2007-2015), the Integrated Management of Childhood Illness (IMCI), National Health Insurance Policy, the Free Maternal Delivery Services [8] and the Community-Based Health Planning and Services (CHPS) Policy 13, 24, 25]. A plethora of non-Governmental and religious Organizations (NGOs) are also into various dimensions of “women’s empowerment” especially in education, health, and economic emancipation.

However, it is not clear the extent to which gender designed programs and policies have impacted on women ability to make decisions and act appropriately regarding their health and the health of their children. For instance, are women in Ghana able to make decisions about their own health or that of their children without censorship by their spouses and other significant family members? Are children born to women with greater autonomy more likely to survive compared to those born to women with weak or no autonomy? Addressing these important questions will contribute to knowledge and adoption of policies and strategies to reduce child mortality and accelerate progress towards the attainment of the SDG3 target 3.2 in Ghana. This study examined the association between women’s autonomy and neonatal, infant and under-five mortality in the Upper East Region of Ghana; a region characterized by patriarchal norms and male dominance.

## Context of the study data

### Methods

Data for this study comes from the Ghana Essential Health Intervention Programme (GEHIP) which was implemented in the Upper East Region of Ghana [26]. GEHIP was implemented to address bottlenecks in Ghana’s flagship primary healthcare Programme known as the Community-based Health Planning and Services (CHPS). As a policy, CHPS is designed to provide a wide range of essential preventive and curative services towards achieving Universal Health Coverage (UHC). The policy is the product of research findings of the “Navrongo Community Health and Family Planning” project which demonstrated significant impact on fertility and child survival [28]

### Ethics Review and Approval

This study was reviewed and approved by the Navrongo Health Research Centre Institutional Review Board; reference number **NHRCIRB127.** Participation in the study was voluntary and written informed consent was obtained from eligible respondent before participation. Consent for non-literate respondents was administered in the preferred local dialect of the study participants and witnessed by an adult literate family or community member. Participants who consented to participate in the survey thumb printed the consent forms and were given copies for their personal keep.

### Sampling and scope of data collected

Data collection involved a cross-sectional randomized cluster household survey among women aged 15-49 years old. First, 66 Enumeration Areas (EAs) proportional to population size were randomly sampled [8]. Household listings for all households in the sampled EAs was then carried out. Households with eligible respondents (women 15-49 years old) in the sampled EAs were then randomly selected proportional to the population size of the Enumeration Area. All eligible women in the sampled households who voluntarily agreed to participate were interviewed. Data was collected over the period 2^nd^ October 2014 to 31^st^ December 2015. The paperless “Open Data Kit” (ODK) software was used in collecting the data [28]. Out of a the 5, 914 interviewed yielding a 76% achieved sample, the analysis was restricted to 3,243 who reported ever having given birth prior to the survey.

Information on detailed reproductive profiles of each woman such as the number of biological sons and daughters alive, at the time of the survey, the sex of the child, date of birth, place of birth, and whether the birth was single or multiple were collected. In this study, a live birth was defined as one in which the child cried or showed signs of life at birth such as pulsation of the umbilical cord or definite muscle movement. For all children who died, information on the age at death was obtained. This information enabled us to generate neonatal, child and under five categories of death. Information on household interactions including decision-making with respect to the use of money and household resources, utilization of health services, sexual coercion and social mobility was also obtained from the study participants. Participants were asked to indicate who usually makes decisions about major household purchases; how money they earn is spent; and purchases for daily needs. Participants were also asked if they can visit families and friends without obtaining permission; refuse to have sex with their partner without any severe consequences and lastly whether they needed permission to seek care at a health facility. These set of questions enabled us to construct a “women’s autonomy” index as the explanatory variable.

### Description of Variables

#### Outcome variables

##### Neonatal mortality

Refers to deaths of newborn children within the first 28 days of life.

##### Infant and under-five mortality

Infant mortality refers to all deaths between birth and the first one-year of life while under-five mortality refers to all deaths between birth and a child’s fifth birthday.

#### Exposure variable

Women’s autonomy: This was generated as an index based on a Principal Component Analysis using five different dimensions of women’s household decision-making with respect to use of money and household resources, utilization of health services, sexual coercion, and social mobility.

Other covariates were: 15-24, 25-34 and 35-49 years; level of education attained categorized as None, Primary/Middle/JHS, and Secondary or above; religious affiliation categorized as Christians, Traditional, Islam and No religion; socio-economic status classified into quintiles (poorest, very poor, poor, less poor and the least poor); and place of residence grouped into three as (Urban, Peri-urban, and Rural). Marital status was classified as Married, Separated, never married, Cohabiting/living together and district of residence.

### Data analysis

The data was analyzed using Stata version 13. Because of our interest in child mortality, the analyses were restricted to women who had ever given birth at the time of the survey.

Appropriate sample weights were applied to account for the cluster sampling design effects. Both descriptive and inferential statistics were used to obtained study results.

#### Women autonomy index

A principal component analysis (PCA) was used to construct an autonomy index for women based on six household decision-making variables; spending decider, decider of daily purchases, decider of major household purchases, can refuse sex, permission to go to health facility, and visits to family and friends. All six components explain 100% of the variation in the data and were also found to be associated with the outcome variable. We computed the statistical adequacy for each variable used for the PCA analysis using the Kaiser-Meyer-Olkin (KMO). The obtained test value ≥0.5 indicated that, all variables were significantly adequately sampled in the analysis generating women’s autonomy. The autonomy index obtained was categorized as least, moderate, and high autonomy.

The index so obtained was used as the exposure variable while the outcome variables were neonatal, infant and under-five mortality. All categorical variables were presented as proportions. First, associations between categorical variables were tested using Pearson’s Chi-squared test. The associations with each of the covariates were then run in univariate logistic regression models and covariates which were significantly associated with women’s autonomy at the 5% level of significance retained in the multivariate logistic model including all other variables a prior from literature. All tests in the inferential analyses were deemed statistically significant at 5% level of statistical significance and a full suite of diagnostic analyses of the models fitted were performed to assess their appropriateness.

## Results

All the background characteristics (age group, highest level of education attained, religious affiliation, socio-economic status, marital status, and place of residence) were highly significantly associated with women’s autonomy status (P≤0.001), Table 1.

**Table 1.**
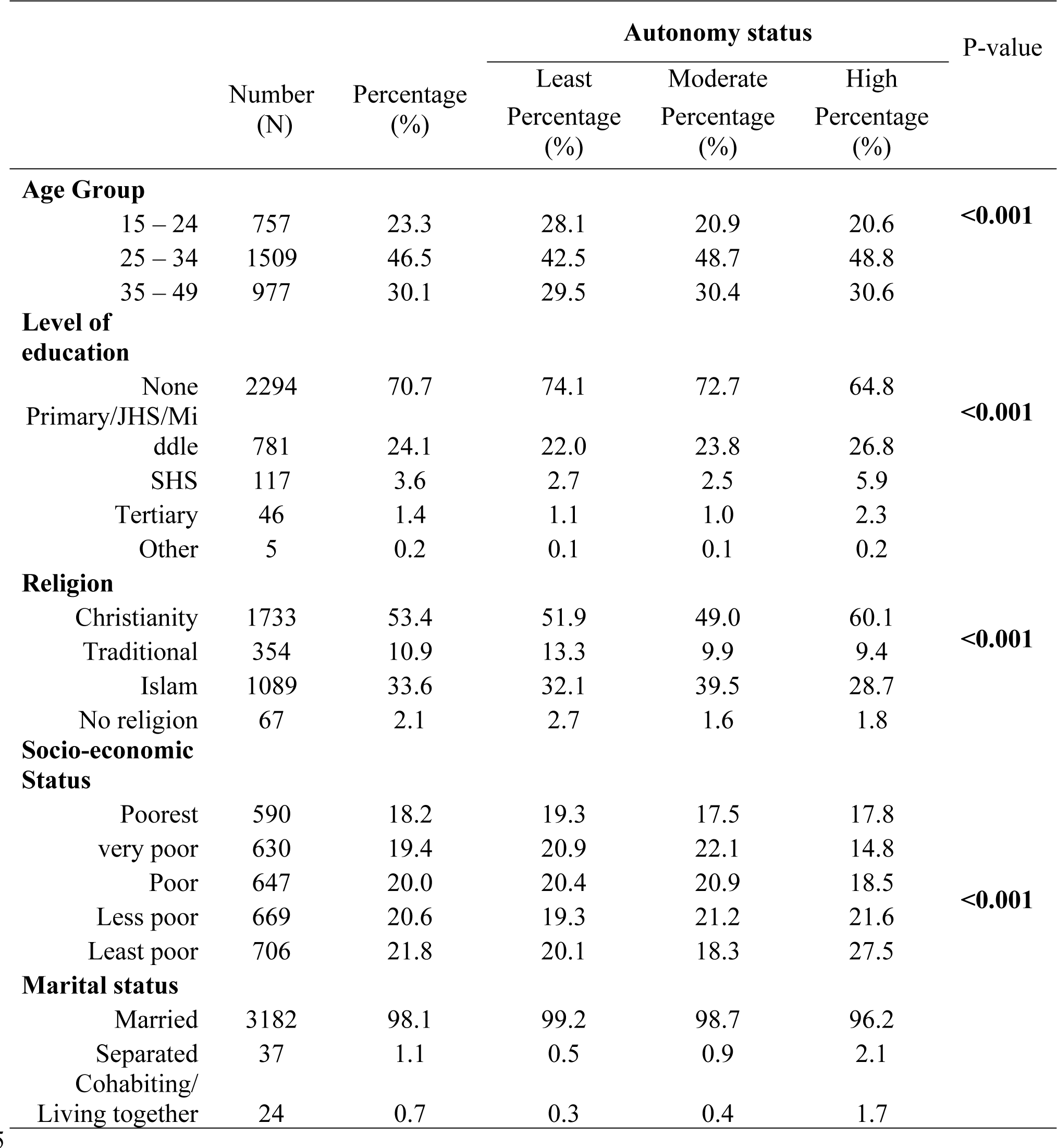
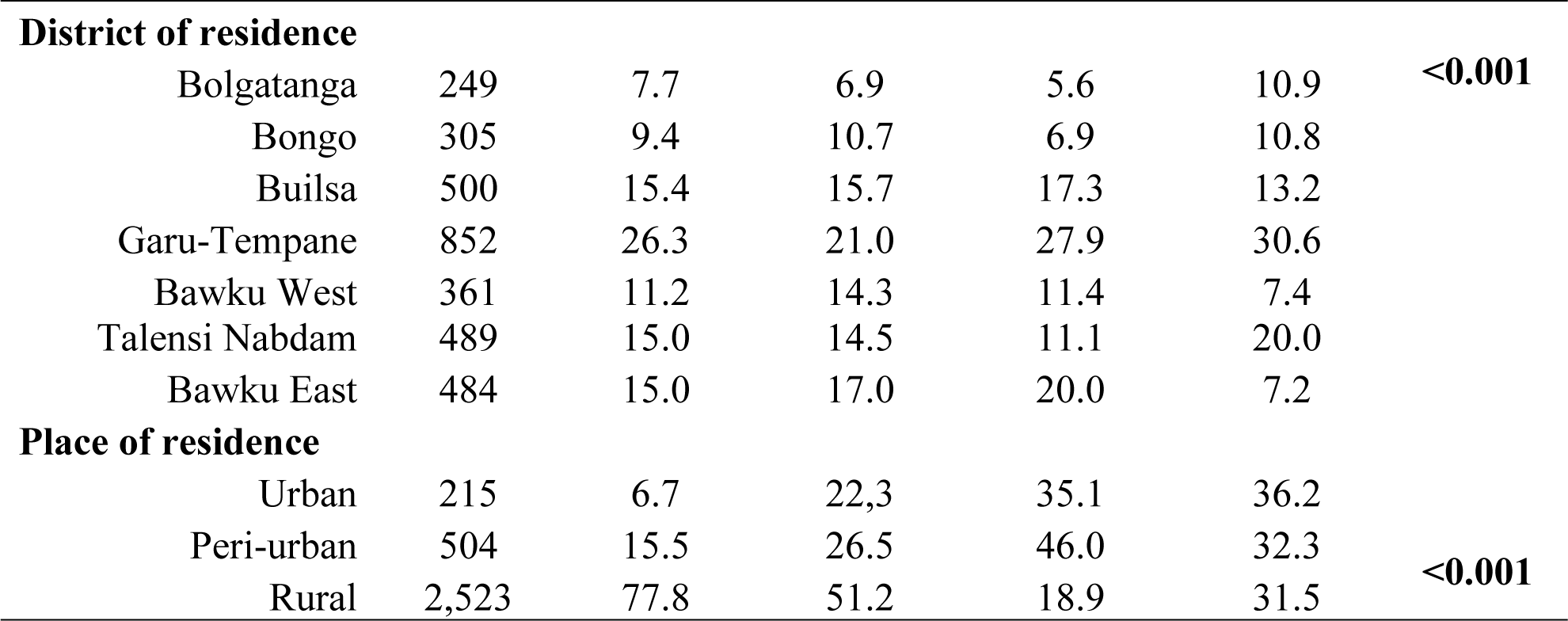
Background characteristics of study participants by autonomy status.

|  | Number<br>(N) | Percentage<br>(%) | Autonomy status |  |  | P-value |
| --- | --- | --- | --- | --- | --- | --- |
|  |  |  | Least<br>Percentage<br>(%) | Moderate<br>Percentage<br>(%) | High<br>Percentage<br>(%) |  |
| **Age Group** |  |  |  |  |  |  |
| 15 – 24 | 757 | 23.3 | 28.1 | 20.9 | 20.6 | **<0.001** |
| 25 – 34 | 1509 | 46.5 | 42.5 | 48.7 | 48.8 |  |
| 35 – 49 | 977 | 30.1 | 29.5 | 30.4 | 30.6 |  |
| **Level of education** |  |  |  |  |  |  |
| None | 2294 | 70.7 | 74.1 | 72.7 | 64.8 | **<0.001** |
| Primary/JHS/Middle | 781 | 24.1 | 22.0 | 23.8 | 26.8 |  |
| SHS | 117 | 3.6 | 2.7 | 2.5 | 5.9 |  |
| Tertiary | 46 | 1.4 | 1.1 | 1.0 | 2.3 |  |
| Other | 5 | 0.2 | 0.1 | 0.1 | 0.2 |  |
| **Religion** |  |  |  |  |  |  |
| Christianity | 1733 | 53.4 | 51.9 | 49.0 | 60.1 | **<0.001** |
| Traditional | 354 | 10.9 | 13.3 | 9.9 | 9.4 |  |
| Islam | 1089 | 33.6 | 32.1 | 39.5 | 28.7 |  |
| No religion | 67 | 2.1 | 2.7 | 1.6 | 1.8 |  |
| **Socio-economic Status** |  |  |  |  |  |  |
| Poorest | 590 | 18.2 | 19.3 | 17.5 | 17.8 | **<0.001** |
| very poor | 630 | 19.4 | 20.9 | 22.1 | 14.8 |  |
| Poor | 647 | 20.0 | 20.4 | 20.9 | 18.5 |  |
| Less poor | 669 | 20.6 | 19.3 | 21.2 | 21.6 |  |
| Least poor | 706 | 21.8 | 20.1 | 18.3 | 27.5 |  |
| **Marital status** |  |  |  |  |  |  |
| Married | 3182 | 98.1 | 99.2 | 98.7 | 96.2 |  |
| Separated | 37 | 1.1 | 0.5 | 0.9 | 2.1 |  |
| Cohabiting/<br>Living together | 24 | 0.7 | 0.3 | 0.4 | 1.7 |  |

| **District of residence** |  |  |  |  |  |  |
| --- | --- | --- | --- | --- | --- | --- |
| Bolgatanga | 249 | 7.7 | 6.9 | 5.6 | 10.9 | <0.001 |
| Bongo | 305 | 9.4 | 10.7 | 6.9 | 10.8 |  |
| Builsa | 500 | 15.4 | 15.7 | 17.3 | 13.2 |  |
| Garu-Tempane | 852 | 26.3 | 21.0 | 27.9 | 30.6 |  |
| Bawku West | 361 | 11.2 | 14.3 | 11.4 | 7.4 |  |
| Talensi Nabdam | 489 | 15.0 | 14.5 | 11.1 | 20.0 |  |
| Bawku East | 484 | 15.0 | 17.0 | 20.0 | 7.2 |  |
| **Place of residence** |  |  |  |  |  |  |
| Urban | 215 | 6.7 | 22.3 | 35.1 | 36.2 | <0.001 |
| Peri-urban | 504 | 15.5 | 26.5 | 46.0 | 32.3 |  |
| Rural | 2,523 | 77.8 | 51.2 | 18.9 | 31.5 |  |

### Distribution of neonatal, child and under-five mortality by women’s autonomy status

Figure 1 shows that proportionally, the highest neonatal (37.3%), infants (37.2%) and under-five (35.5%) deaths occurred among women with higher autonomy. The higher the maternal autonomy status, the greater the proportion of neonatal deaths. Maternal autonomy was significantly associated with neonatal, infant and under-five mortality.

**Fig 1.**
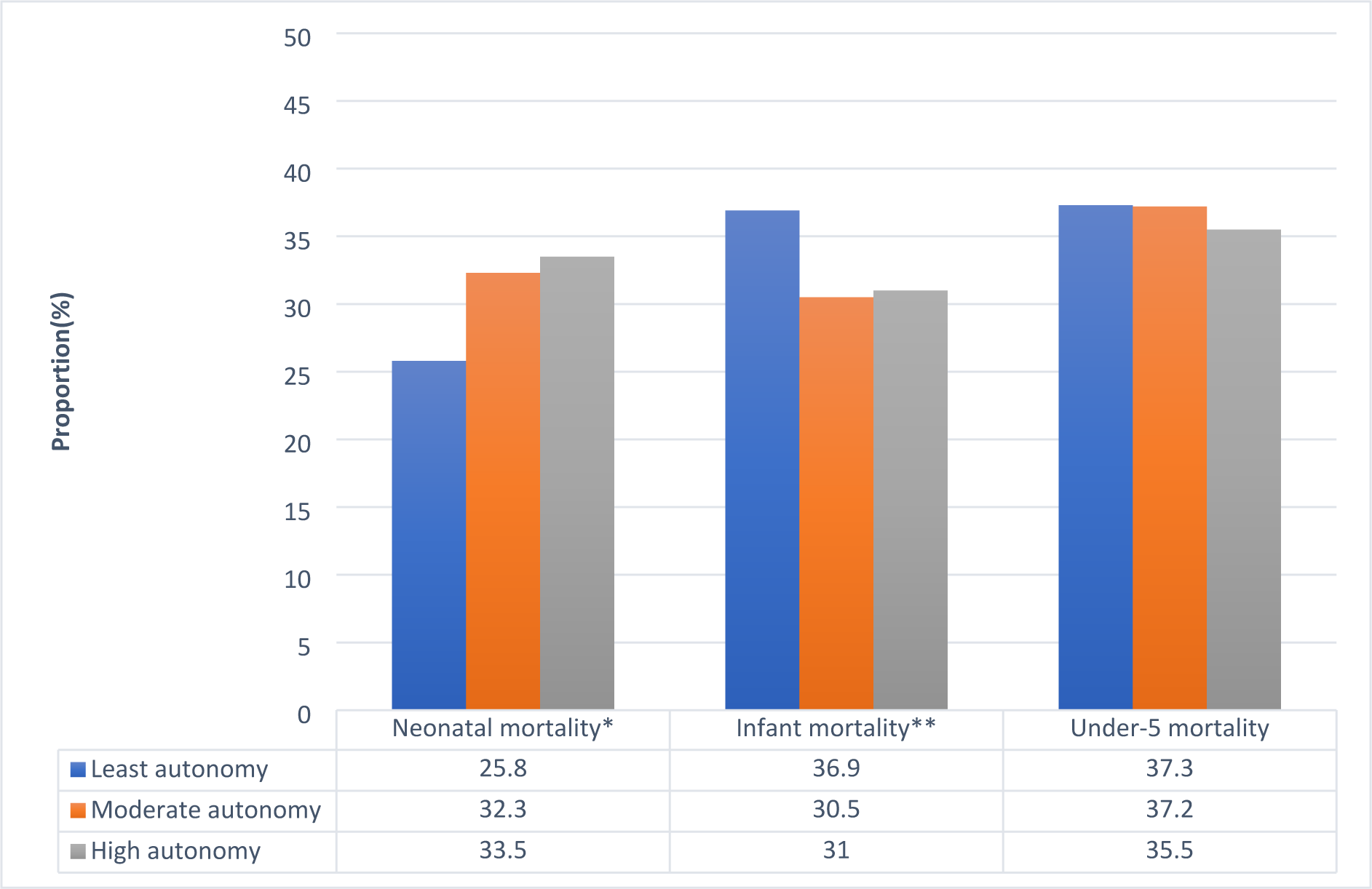
Percentage distribution of neonatal, infant and under-five mortality by women’s autonomy status. Level of significance; ***p<0.001; **p<0.01 and *p<0.05

Presented in table 2 is the bivariate and multivariate regression analysis of the association of women’s autonomy with neonatal, infant and under-five mortality. Paradoxically, we found a counterintuitive relationship between women’s autonomy and neonatal, infant and under-five mortality. In the unadjusted regression model, moderate women’s autonomy was associated with a statistically significant increased risk of neonatal death (OR: 1.61, 95% CI 1.03-2.51) compared to those born to women with least autonomy. In the adjusted regression model, the risk of neonatal death among women with moderate autonomy compared to those with least autonomy increased slightly (aOR: 1.76, 95% CI 1.07- 2.89) and remained statistically significant. Similarly, higher women’s autonomy was significantly associated with increased neonatal mortality even after adjusting for socio- demographic factors (aOR: 1.75, 95% CI, 1.04-2.93).

**Table 2.** Bivariate and Multivariate analysis of the Association between women’s autonomy and neonatal, infant and under-five mortality.

|  | Neonatal mortality |  | Infant mortality |  | Under-five mortality |  |
| --- | --- | --- | --- | --- | --- | --- |
| Variable | Bivariate | Multivariate | Bivariate | Multivariate | Bivariate | Multivariate |
|  | OR [95% CI] | aOR [95% CI] | OR[95% CI] | aOR [95% CI] | OR[95% CI] | aOR [95% CI] |
| Autonomy |  |  |  |  |  |  |
| Least | 1 | 1 | 1 | 1 | 1 | 1 |
| Moderate | 1.61[1.03,2.51]* | 1.76[1.07,2.89]* | 1.04[0.78,1.38] | 1.01[0.75,1.36] | 0.95[0.79,1.15] | 0.95[0.78,1.15] |
| High | 1.99[1.30,3.05]** | 1.75[1.04,1.74]* | 1.36[1.07,1.74]* | 1.31[0.93,1.86] | 1.20[0.95,1.86] | 1.15[0.89,1.47] |
| Socio-demographic Characteristics |  |  |  |  |  |  |
| Level of education attained |  |  |  |  |  |  |
| None | 1 | 1 | 1 | 1 | 1 |  |
| Prim/JHS/Middle | 0.53[0.31,0.93]* | 0.83[0.42,1.61] | 0.34[0.23,0.49]*** | 0.94[0.63,1.40] | 0.40[0.31,0.53]*** | 0.87[0.65,1.15] |
| SHS + | 0.75[0.35,4.21] | 0.70[0.16,3.13] | 0.21[0.11,0.40]*** | 0.39[0.16,0.94]* | 0.24[0.16,0.58]*** | 0.39[0.18,0.87]* |
| Age Group |  |  |  |  |  |  |
| 15-24 | 1 | 1 | 1 | 1 | 1 | 1 |
| 25-34 | 1.58[0.71,3.50] | 1.50[0.60,3.73] | 3.13[2.01,4.87]*** | 3.14[1.94,5.07]*** | 2.50[1.74,3.59]*** | 2.38[1.62,3.50]*** |
| 35-49 | 4.51[2.32,8.79]*** | 3.97[1.93,8.13]** | 20.55[13.13,32.17]*** | 17.17[10.53,28.02]*** | 13.41[9.48,18.99]*** | 10.87[7.46,15.84]*** |
| Religious<br>Affiliation |  |  |  |  |  |  |
| Christianity | 1 | 1 | 1 | 1 | 1 | 1 |
| Traditional | 1.68[1.14,2.48]* | 1.15[0.70,1.89] | 2.27[1.70,3.03]*** | 1.11[0.80,1.54] | 1.88[1.46,2.43]*** | 1.27[1.01,1.60]* |
| Islam | 0.60[0.36,1.01] | 0.63[0.34,1.18] | 0.66[0.51,0.86]** | 0.63[0.46,0.88]** | 0.68[0.55,0.84]*** | 0.69[0.55,0.86]** |
| No religion | 0.68[0.23,2.04] | 0.57[0.19,1.68] | 1.62[0.87,3.00] | 1.71[1.13,2.59]* | 1.71[1.13,2.59]* | 1.29[0.83,2.01] |
| Socio-economic<br>status |  |  |  |  |  |  |
| Poorest | 1 | 1 | 1 | 1 | 1 | 1 |
| Very poor | 1.10[0.56,2.14] | 0.97[0.47,2.00] | 0.85[0.59,1.22] | 0.93[0.64,1.36] | 0.97[0.78,1.20] | 1.17[0.94,1.47] |
| Poor | 0.75[0.42,1.33] | 0.99[0.52,1.88] | 0.81[0.54,1.21] | 1.31[0.81,2.09] | 0.82[0.66,1.03] | 1.04[0.83,1.31] |
| Less poor | 0.73[0.42,1.26] | 0.74[0.36,1.50] | 0.93[0.60,1.44] | 1.11[0.70,1.75] | 0.02[0.77,1.35] | 1.19[0.86,1.65] |
| Least poor | 0.72[0.42,1.25] | 0.86[0.46,1.59] | 0.74[0.49,1.10] | 1.20[0.76,1.89] | 0.82[0.62,1.08] | 1.09[0.85,1.41] |
| Place of Residence |  |  |  |  |  |  |
| Urban | 1 | 1 | 1 | 1 | 1 | 1 |
| Semi-urban | 0.42[0.20,0.87]* | 0.32[0.13,0.81]* | 0.78[0.24,1.44] | 0.93[0.50,1.74] | 0.68[0.44,1.04] | 0.69[0.48, 1.01] |
| Rural | 0.51[0.27,0.97]* | 0.38[0.19,0.75]** | 0.70[0.45,1.07] | 0.68[0.43,1.08] | 0.69[0.52,0.92*] | 0.63[0.48,0.083]** |

Level of significance; \*\*\*p<0.001; \*\*p<0.01 and \*p<0.05

Women’s autonomy was statistically significantly associated with infant mortality in the unadjusted regression model (OR=1.36, 95% CI=1.07-1.74) but not in the adjusted model (aOR=1.31, 95% CI= 0.93-1.86). Similarly, women’s autonomy was not significantly associated with under-five mortality. However, in both the unadjusted and adjusted regression models, women with high autonomy were 20.0% and 15.0% respectfully more likely to have experienced an under-five death when compared with those with least autonomy.

## Discussion

In this study, we examined the association between women’s autonomy and neonatal, infant and under-five mortality in the Upper East Region of Ghana s study. This is important because the quality of care that children receive is greatly depended on their household circumstances. As primary caregivers, mothers play a critical role in making decisions that directly or indirectly affects the health of children. Yet, the extent to which constricted women’s autonomy adversely affects child mortality has not been sufficiently interrogated in Ghana. Our findings revealed a linear relationship between neonatal mortality and women’s autonomy status; with the highest neonatal deaths occurring among women with high autonomy status. In contrast, the highest infants and under-five deaths occurred among women with the least autonomy status. The adjusted region analysis showed that maternal autonomy was significantly associated with neonatal, infant and under-five mortality. Compared to women with the least autonomy status, infants of those with moderate autonomy and high autonomy were significantly more likely to die.

Our findings differ from some previous studies involving multiple African countries and other developing countries [21, 29, 30] . Adedini and colleagues whose study involved 18 African countries including Ghana, found that higher position of women in a household (based on a composite score of marital dyads), was significantly associated with lower risk of neonatal mortality [29]. Memiah and colleagues’ multi-country study in East Africa (Burundi, Kenya, Rwanda, Tanzania, and Uganda) found that women’s ability to exercise discretion not to have sex without suffering any consequences was significantly associated with 16% less likelihood of under-five mortality. Similarly, Doku and colleagues’ meta-analysis of pooled data from 59, “Low- and Middle-Income Countries” (LMICs), found that women with a lower score of Individual-level Women’s Empowerment Index (ILWEI) had 18%, 12% and 17% higher risk of neonatal, infant and under-five deaths respectfully than those with higher ILWEI. However, country level analysis revealed an inconsistent relationship between WEI and mortality outcomes [21].

The evidence from this study is consistent with a few studies [31]. that showed a weak or counter intuitive statistically significant associations between women’s autonomy and maternal healthcare utilization, higher maternal autonomy is generally associated with higher utilization of antennal care services, skilled birth delivery, postnatal and general child healthcare [13, 15, 17–20]; which leads to better child health outcomes. This notwithstanding, our results suggest that the role of significant others in decision-making in child health during the neonatal and infant period remains critical in this study setting. The observed differences reported by the various studies may be due the differences in the variables used to measure women’s autonomy index. Thus, there is need to re-examine the appropriateness of autonomy models that focus on individualism. This is critical given the imbedded context of social relations and the role of the family in childcare in this and similar study context in Ghana. Contextual models that incorporates the role of significant others in this and similar settings in Africa may offer better child health outcomes.

Our results do not imply that women empowerment and autonomy is not important in attaining SDG3. Our study clearly demonstrated the positive effect of maternal educational attainment in child mortality. In the adjusted regression model, maternal education remained significantly associated with infant and under-five mortality. Attaining secondary or higher education was significantly associated with a decreased risk of children dying before attaining their first date of birth or fifth birthday respectfully when compared with children of women who had no education. In fact, women who attained Primary/JHS/Middle education were 60% less likely to experience an under-five death compared to those with no education. These findings are consistent with empirical literature in Ghana and other countries in Sub-Saharan Africa [32–38]. and shows that women empowerment remains an important goal and, a means to attaining significant reductions in child mortality in Ghana and Africa. For instance, Babayara et al. [33] used data from the “Navrongo Demographic and Surveillance system” and found that mothers who no education accounted for 82.2% of under-five deaths.

We conclude that in this study setting women’s autonomy was not protective of child mortality. In fact, compared to women with diminished autonomy, those with higher autonomy were significantly more likely to experience neonatal and infant mortality. Given the imbedded context of social relations and the family system in the study context and similar settings in Ghana, there is need to re-examine the appropriateness of autonomy models that focus on individualism. Contextual models that factor in the unique African family systems are more likely to yield better outcomes.

## Limitations and strengths of the Study

This study has some inherent limitations. First, it relied on a cross sectional household survey data and hence is unable to establish any causality between “women’s autonomy” and the study outcomes (neonatal, infant and under-five mortality). Secondly, “women’s autonomy” is a fluid and multifaceted concept which is difficult to measure accurately. The indirect approach used in eliciting information to generate the autonomy variable may not be very accurate. Notwithstanding these few limitations, this study contributes immensely to knowledge regarding women’s autonomy and child health and support earlier calls for prioritizing family and community level factors that promote increased utilization of tailored essential maternal and new-born care interventions such as appropriate breastfeeding practices, skilled birth attendance, timely vaccinations, vitamins and minerals supplementation and prophylactic antibiotics as recommended by the WHO as a strategy to accelerating progress towards attainment of SDG 3 in Ghana.

## Acknowledgement

We wish to acknowledge and appreciate the study participants for making time out of their busy schedules to participate in the study and for their honest and sincere responses to questions asked which contributed greatly to the quality of the data. We equally acknowledge the contribution of the data collectors for their invaluable contribution. Finally, we appreciate the Management and Staff of the Navrongo Health Research Centre and the Department of Population and Health, of the University of Cape for their various support during the study.

## Data Availability

Data are available for public use and can be obtained by request to the Director of Navrongo Health Research Centre, Box 114, Navrongo

